# Little Risk of the COVID-19 Resurgence on Students in China (outside Hubei) Caused by School Reopening

**DOI:** 10.1101/2020.04.04.20053645

**Authors:** Cheng Long, Tieyong Zeng, Xinmiao Fu

## Abstract

**Objective:** School reopening has not yet started in China where the COVID-19 outbreak has reached ending stage, largely due to a great concern about COVID-19 infections on students. We attempted to quantitatively evaluate the risk of COVID-19 infections on students caused by school reopening.

**Study design:** We collected the data of the numbers of teachers, population size and newly confirmed COVID-19 cases in the past 14 days in typical provinces/cities of China, and then analyzed the risk of COVID-19 infections in schools with respect to each province/city.

**Methods:** A step-by-step procedure was explored to calculate the probability of COVID-19 infections on students as transmitted from infected teachers. Two critical assumptions for analysis were proposed: (i) only locally generated cases were counted while imported cases were omitted; (ii) the secondary attack rate of the COVID-19 virus in schools is similar to that in households in China, ranging from 3-10%.

**Results:** The probability of COVID-19 resurgence within one week on students of primary, middle and high schools in China (outside Hubei) is extremely low (<0.2%) in each province/city, and such probability can be updated daily and weekly based on the newly confirmed cases in the past 14 days. In some areas without newly confirmed cases in the past 14 days, the risk is zero.

**Conclusions:** Our work provides guidance for local governments to make risk level-based policies for school reopening. Currently, the risk of COVID-19 infections on students is extremely low in China (outside Hubei) and therefore school reopening can be initiated without the endanger of infections on students.

---

The novel coronavirus diseases (COVID-19) outbreak is going on in China and has resulted in 80000 confirmed cases and over 3100 deaths as of 10 March 2020 ^1^. Since March 11, China only reported 20 or less new cases, most of which are concentrated in Hubei Province and its capital Wuhan City, the epicenter of the outbreak. As such, China (outside Hubei) has entered a new stage of epidemic prevention and control coupled with a stepwise restoration of social and economic operations ^2^. In particular, it is highly demanding to reopen schools because the delay of schooling time for approximately two months has substantially impacted on more than 100 millions of families in China. Nevertheless, none of schools has reopened so far across the country, largely due to a great concern about the risk of COVID-19 infection on children ^3^. Here we show by statistical probability analysis that the risk of COVID-19 resurgence caused by school reopening is negligible.

Our analysis is based on several assumptions as follows. First, a period of the past 14 days was set as a reference for risk assessment, given that the incubation period of COVID-19 ranges from 1-14 days with a mean of 5-6 days ^2^. Second, the probability of infection in the coming week is proportional to the number of newly confirmed COVID-19 cases in the past 14 days. Third, only locally generated cases are counted while imported cases are omitted (Note: all travelers entering China are required a quarantine for 14 days ^4^). Forth, only primary, middle and high schools were analyzed while colleges and universities were excluded because their students are from across the country (including Hubei Province and Wuhan City) but not solely locally living. Fifth, we assume that all students are healthy, given that only 0.9% of over 50000 COVDI-19 cases in China are aged 0-9 years and 1.2% are 10-19 years ^5^; as such, we simply focus on the potential transmission from teachers to students (or between teachers), excluding that from students to students or teachers. Last, we assume the secondary attack rate of the COVID-19 virus in schools is similar to that in households, ranging from 3-10% ^2^.

Under the above assumptions, we collected the data of population size, number of teachers and new COVID-19 cases in the past 14 days in typical provinces/cities (refer to **Table S1**) that have been most affected by the outbreaks and/or are most economically important in China. We then estimated the probability of COVID-19 transmission step by step, as detailed in **Table S2**. Specifically, we first calculated the probability that at least one teacher has been infected and then estimated the probability of the infected teacher(s) to students or teachers (for detail, refer to **Table S2**).

Results show that the probability in all areas in the coming week (from 13-19 March) is extremely low (except Beijing), ranging from 0.01%-0.13%. If the number of new confirmed cases in the past 14 days is zero (e.g., Henan, Zhejiang, Jiangxi, Anhui, Guangzhou and Shenzhen), then the risk is zero. If new cases as of 9 March were counted, then the probability of COVID-19 resurgence from 10-16 March would be a slightly higher, ranging from 0.01-0.37%. The probability for Beijing is highest because of 10 new cases reported on 26 March (refer to **Table S1**).

In summary, our analyses suggest that the probability of COVID-19 resurgence regarding school reopening is low in all provinces/cities outside Hubei (all <0.5%). Such probability can be updated daily or weekly based on the number of new cases in the past 14 days. Furthermore, the overall endanger of COVID-19 infection in students would be extremely low from the clinical point of view, given that in China only a very small proportion of the COVID-19 cases aged under 19 years have developed severe (2.5%) or critical disease (0.2%) and that among a total of 1023 deaths only one death was from this age group, as revealed by earlier reports of China CDC and WHO ^2, 3, 5^. In addition, daily temperature monitoring on teachers and inspecting their body status are necessary, and anyone who has symptoms of fever and cough should be immediately isolated away from schools and subject to further clinical diagnosis.

Our work may provide guidance for provincial governments to make risk level-based, differentiated control measures, by which societal activities, particularly school reopening, are effectively restored and the potential risk of COVID-19 resurgence is strictly controlled. During this process, governments always get ready to immediately react to any new COVID-19 cases or clusters. Furthermore, if parents raise deep concerns about the risk of infections on student by potentially infected teachers given the above control measures, one supplementary strategy that can further reduce the risk is to screen all the teachers with COVID testing kits to identify potentially infected ones. Meanwhile, all the teachers should be informed to avoid any unnecessary clustering activities that might make them to be potentially infected.

**Table 1.** Probability of COVID-19 resurgence after school reopening^*a*^

| Province /cities | Total cases | Population (10<sup>4</sup>) | No. of teachers (10<sup>4</sup>) | 13-19 March 2020 |  | 10-16 March 2020 |  |
| --- | --- | --- | --- | --- | --- | --- | --- |
|  |  |  |  | New cases <sup>b</sup> | Probability (%) <sup>c</sup> | New cases <sup>b</sup> | Probability (%) <sup>c</sup> |
| Guangdong | 1353 | 11346 | 96.7 | 2 | 0.03-0.08 | 3 | 0.04-0.13 |
| Henan | 1272 | 9605 | 99.3 | 0 | 0 | 1 | 0.02-0.05 |
| Zhejiang | 1215 | 5737 | 40.8 | 0 | 0 | 0 | 0 |
| Hunan | 1018 | 6898 | 53.0 | 1 | 0.01-0.04 | 2 | 0.02-0.08 |
| Jiangxi | 935 | 4647 | 42.2 | 0 | 0 | 1 | 0.01-0.05 |
| Anhui | 990 | 6323 | 48.7 | 0 | 0 | 1 | 0.01-0.04 |
| Shandong | 759 | 10047 | 85.1 | 3 | 0.04-0.13 | 2 | 0.03-0.08 |
| Jiangsu | 631 | 8050 | 60.3 | 0 | 0 | 0 | 0 |
| Fujian | 296 | 3973 | 32.5 | 0 | 0 | 2 | 0.02-0.08 |
| Beijing | 429 | 2153 | 12.3 | 3 | 0.03-0.09 | 13 | 0.11-0.37 |
| Shanghai | 344 | 2423 | 11.6 | 1 | 0.01-0.02 | 3 | 0.02-0.07 |
| Guangzhou | 347 | 1490 | 10.1 | 1 | 0.01-0.03 | 1 | 0.03-0.01 |
| Shenzhen | 419 | 1302 | NA <sup>b</sup> | 0 | 0 | 0 | 0 |

<sup>a</sup> Steps for probability calculation are presented in **Table S2**.
<sup>b</sup> Daily new cases are shown in **Table S1**. NA: not available.
<sup>c</sup> The secondary attack rate was set as 3%-10% by referring to the estimates on family clusters <sup>2</sup>.

## Data Availability

All data are included in the manuscript

## Acknowledgments

This work is support by the National Natural Science Foundation of China (No. 31972918 and 31770830 to XF). We declare no competing interests.

